# Enhancing the Diagnostic Accuracy of Amyloid PET: The Impact of MR-Guided PET Reconstruction

**DOI:** 10.1101/2025.01.04.25319996

**Authors:** M. Mehdi Khalighi, Christina B. Young, Skylar Weiss, Michael Zeineh, Guido Davidzon, Elizabeth Mormino, Greg Zaharchuk

**Affiliations:** Radiology Department, Stanford University; Neurology Department, Stanford University

**Keywords:** PET/MR, MR Priors, Anatomical Priors, PET Reconstruction, Amyloid

## Abstract

^18^F-Florbetaben (FBB) uptake in the supratentorial cortex is indicative of amyloid positivity. Due to PET’s low spatial resolution, image noise, and spill-over of signal from adjacent white-matter into gray-matter, trained readers may provide inconsistent reads and quantitative calculations like Centiloids (CLs) are also affected. A set of 264 ^18^F-Florbetaben (amyloid) PET/MRI exams were reconstructed using conventional ordered subset expectation maximization (OSEM) method and MR-guided block sequential regularized expectation maximization (MRgBSREM) method. Images from 264 patients reconstructed by OSEM method and rated by 3 trained readers. Fifty-three exams were rated inconsistently and were mixed with another 53 exams which were rated consistently. These 106 subjects were then rated by our readers using the MRgBSREM PET reconstruction method. CLs were measured using both reconstruction methods. There is significant correlation between CL measured by OSEM and MRgBSREM methods with R2=0.99. The number of inconsistent exams dropped by 64% using MRgBSREM method as compared with OSEM method. Using Fleiss-Kappa statistical test, the agreement between readers was raised from “Fair” to “Significant” in the 106-subjects subset. PET reconstruction with MR priors can significantly improve the consistency of ratings among trained readers. Given the prevalence of inconsistent ratings in amyloid PET, methods that enhance the ability to distinguish intermediate amyloid levels could be valuable for the widespread adoption of this modality.

## I. INTRODUCTION

Alzheimer’S disease (AD) is characterized by the accumulation of extracellular amyloid-beta (Aβ) plaques and intracellular tau tangles in the brain. Positron Emission Tomography (PET) imaging has emerged as a valuable tool for visualizing and quantifying these pathological hallmarks, aiding in diagnosis, staging, and monitoring of AD [1,2]. Simultaneous PET and MRI (PET/MRI) modality provides a single combined imaging examination for a vulnerable patient population and improves the accuracy of diagnostics by providing high resolution structural MRI images combined with relatively lower resolution PET images [3,4].

In clinical settings, the amyloid PET images are rated by trained readers and are typically interpreted as either positive or negative; however, a portion of scans fall into an intermediate range, leading to inconsistent ratings among trained readers. Another approach for examining amyloid PET images is the use of the Centiloid (CL) scale, a common quantitative approach that yields a continuous measure of amyloid burden [5]. However, both of these approaches are impacted by PET’s low spatial resolution, which can result in spill-in effects from adjacent white matter to gray matter or spill-over effects from gray matter to adjacent tissues. Additionally, variability can arise from different acquisition windows as the signal increases throughout the scan [6], as well as from other factors such as image noise or atrophy with a thinned cortex.. Furthermore, the CL scale measures uptake in the global cortex and lacks sensitivity to local uptake, which is crucial for early detection.

To address these limitations, there is a growing interest in developing techniques to improve the spatial resolution and image quality of PET images [7,8]. Higher resolution images can enhance both quantitative accuracy and visual interpretation. Additionally, reducing scan time or radiation dose, while maintaining image quality, is crucial for patient comfort and safety, especially for those undergoing multiple scans.

Recent advancements in PET/MR imaging have enabled the reconstruction of high-resolution PET images with 1 mm^3^ isotropic resolution using MR anatomical priors [9]. This novel approach has the potential to significantly improve the accuracy and consistency of amyloid PET imaging. Our research aims to explore the impact of high-resolution PET images on both quantitative and qualitative assessments of amyloid burden, ultimately contributing to more accurate and reliable diagnosis and monitoring of AD.

## II Methods

Two hundred sixty-four participants provided informed consent under a Stanford institutional review board (IRB) protocol. Each subject underwent a 40-minute exam on a simultaneous PET/MR scanner (SIGNA PET/MR, GE Healthcare). Subjects were injected with 317 ± 32 MBq of ^18^F-Florbetaben and scanned 70 minutes post-injection. High resolution 3D *T*_*1*_-weighted inversion-recovery, fast spoiled gradient recalled echo (IR-FSPGR) images (repetition time = 7.7 ms, echo time = 3.1 ms, inversion time = 400 ms, flip angle = 11, spatial resolution=1.2 × 1.1 × 1.1 mm), and 3D *T*_*2*_ fluid-attenuated inversion recovery (FLAIR) CUBE images (repetition time = 6 s, echo time = 164 ms, inversion time = 1766 ms, spatial resolution=1 mm_3_), were acquired simultaneously using Alzheimer’s disease neuroimaging initiative (ADNI) 3 protocols.

PET images were reconstructed between 90 and 110 minutes after injection using two methods. In the first method, PET data were reconstructed into 5-min frames using conventional ordered subset expectation maximization (OSEM) method and then the 5-min frame data were realigned and summed. The OSEM reconstructions parameters were 28 subsets, 3 iterations, 32 cm field of view (FOV), 256 ⨯ 256 matrix, 2.78 mm slice thickness, an in-plane filter with a 3.5 mm full width half maximum (FWHM) spatial cut-off frequency and an axial Standard z-filter (a finite impulse response, FIR, filter with coefficients [1, 4, 1]). In the second method, PET data were reconstructed using MR-guided block sequential regularized expectation maximization (MRgBSREM) [9] with MRI *T*_*1*_-weighted and *T*_*2*_-weighted images as priors. The MRgBSREM regularization parameters were chosen as β=50 and β_m_=25, and images were reconstructed with 1mm isotropic spatial resolution. MR attenuation correction was applied using zero-TE (ZTE) and Dixon MR imaging [10]. All PET images were reconstructed using the time-of-flight information along with corrections for randoms, dead-time, scatter and point-spread-function. Motion correction using short PET frame registration [11] was applied in MRgBSREM PET reconstruction as detailed in [12]. Preprocessing followed ADNI procedures such that FreeSurfer v7 ROIs from the

Desikan aparc+aseg atlas were defined on each participant’s *T*_*1*_-weighted MRI images; native space FreeSurfer ROIs were used to extract intensity values from amyloid PET images; mean uptake across frontal, cingulate, lateral parietal, and lateral temporal cortical regions were calculated; and standardized uptake value ratios (SUVRs) were calculated using a whole cerebellum reference region. Following procedures described in [5], SUVRs were then converted to Centiloids using the equation [13]: Centiloid = 157.15 x SUVR – 151.87, which is the same equation as ADNI as preprocessing procedures were equivalent. The SNR, calculated as the mean SUVR divided by its standard deviation was obtained in frontal, anterior/posterior cingulate, lateral parietal, and lateral temporal ROIs, using both OSEM and MRgBSREM PET reconstruction methods. The SNR was compared between OSEM and MRgBSREM PET reconstruction methods in each ROI.

Three board-certified radiologists rated the PET images reconstructed by OSEM method for all subjects as either amyloid positive or amyloid negative. Of the 264 subjects, 53 (20%) were rated inconsistently (two out of three readers agreed) and the other 211 subjected were rated consistently by all trained reader as either positive (71 subjects) or negative (140 subjects). To investigate the impact of MRgBSREM PET reconstruction method in a blinded reading process, the 53 inconsistently rated subjects were mixed with 53 consistently rated exams (18 positive cases and 35 negative cases) and then all 106 exams were rated by the same three trained readers using the MRgBSREM method. Additionally, a control group consisting of 10% of all exams (26 exams), where half of them were consistently rated and the other half were inconsistently rated, were randomly selected and re-rated by the three trained readers using the conventional PET reconstruction method (OSEM) to assess intra-rater variability. Inter-rater variability was measured using Fleiss’ kappa for both conventional PET reconstruction and PET reconstruction with MR priors.

## III. Results

Fig. 1(a) shows the age distribution of all 264 subjects with average age of 72 ± 9 years. Forty five percent of all subjects were female. Fig. 1(b) shows that about half of the subjects were cognitively unimpaired (CU) while the other half has been diagnosed with mild cognitive impairment (MCI), dementia or other diseases.

**Fig. 1.**
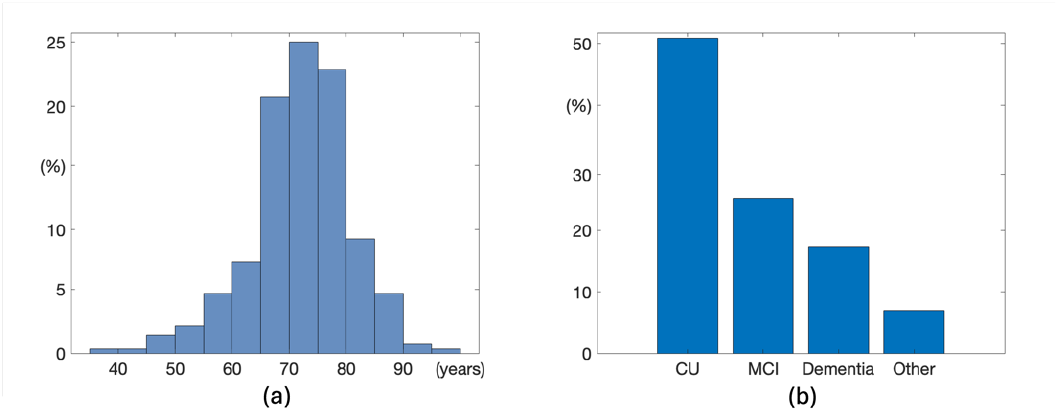
(a) shows the age distribution of all 264 subjects with average age of 72 ± 9 years. (b) shows that about half of the subjects were cognitively unimpaired (CU) while the other half has been diagnosed with mild cognitive impairment (MCI), dementia or other diseases

Fig. 2 illustrates the impact of MR-guided reconstruction on a subject who was initially rated inconsistently by readers using OSEM PET reconstruction (two negative, one positive). When the same subject was evaluated using images reconstructed by MRgBSREM method, all readers consistently rated the scan as negative. The CL decreased from 40 (OSEM) to 21 (MRgBSREM), demonstrating how improved spatial resolution can enhance both visual interpretation and quantitative accuracy of amyloid PET imaging.

**Fig. 2.**
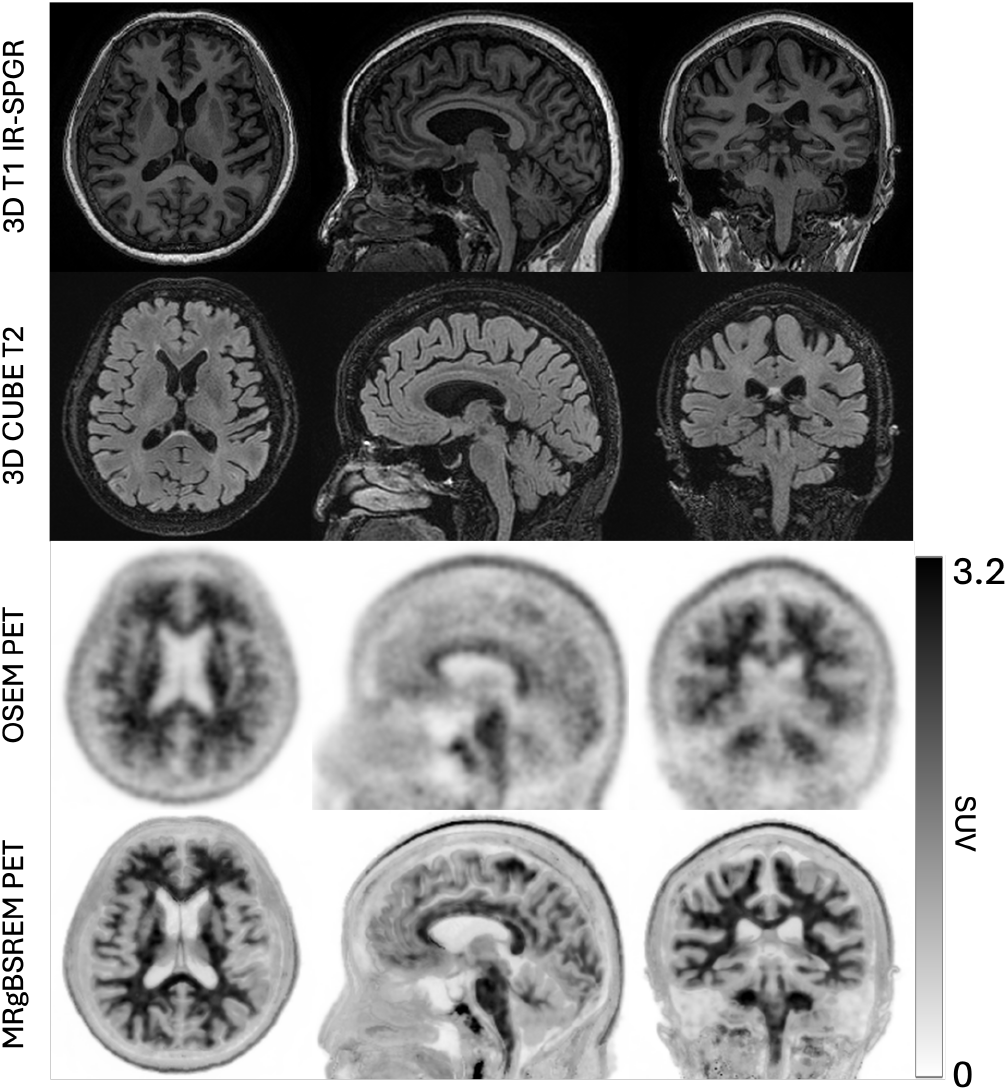
Shows the impact of MR-guided reconstruction on a subject who was initially rated 2 negative and one positive by readers using OSEM method. All readers rated the exam amyloid negative using MRgBSREM method. The CL decreased from 40 (OSEM) to 21 (MRgBSREM), demonstrating how improved spatial resolution can enhance both visual interpretation and quantitative accuracy of amyloid PET imaging.

Fig. 3(a) illustrates the distribution of CL derived from MRgBSREM closely follows that of OSEM PET reconstruction methods using all 264 subjects, confirmed in Fig. 3(b) with a strong correlation between the two methods (R2 = 0.99) and the Bland-Altman plot in Fig. 3(c). This figure shows that subjects with low cortical FBB uptake (CL < 50), may have lower CL when measured with MRgBSREM compared to OSEM method due to the removal of white matter spill-in signal. Conversely, subjects with higher cortical uptake (CL > 50), may show increased CL with MRgBSREM due to the resolution of spill-over signal from gray matter to adjacent tissues.

**Fig. 3.**
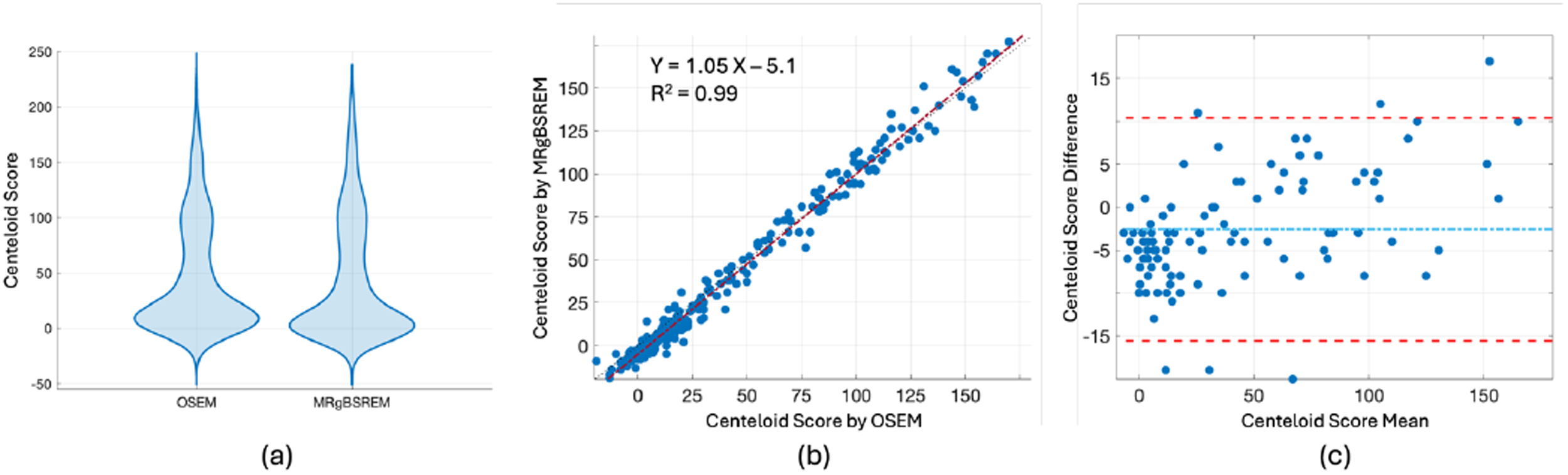
(a) shows the distribution of CL derived from OSEM an MRgBSREM methods. (b) demonstrates a strong correlation between CL derived from the two methods (R2 = 0.99). The Bland-Altman plot in (c) shows the agreement between CL from both methods. It shows that subjects with low cortical FBB uptake may exhibit lower CL when measured with MRgBSREM compared to OSEM method. Conversely, subjects with higher cortical uptake may show increased CL with MRgBSREM.

The SNR, calculated in frontal, anterior/posterior cingulate, lateral parietal, and lateral temporal ROIs for all subjects, was measured at 4.0 ± 0.9 for the OSEM method and 3.5 ± 0.9 for the MRgBSREM PET reconstruction method. Fig. 4 shows a close match between OSEM and MRgBSREM signal-to-noise ratios, with slightly higher SNR for the OSEM method. While OSEM employs a post-processing filter (3.5 mm FWHM spatial cut-off frequency and Standard z-filter) at the end of PET reconstruction, MRgBSREM controls noise through a weighted penalty (β=50) during the iterative process. The MRgBSREM’s SNR can be improved by increasing the β-value, but at the cost of spatial resolution.

**Fig. 4.**
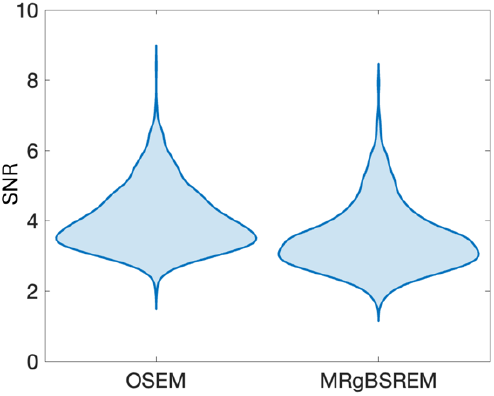
Shows the SNR comparison between the OSEM and MRgBSREM PET reconstruction methods, calculated in frontal, anterior/posterior cingulate, lateral parietal, and lateral temporal ROIs. It shows a close match between the two methods, with slightly higher SNR for the OSEM method, which utilizes a postprocessing filter (3mm spatial cut-off freq and Standard z-filter) compared to the MRgBSREM method (β=50 and βm=25).

Fig. 5 and 6 demonstrate the impact of PET reconstruction with MR priors on reader assessments. In Fig. 5, three subjects initially rated as inconsistent negative (two negative, one positive rating) using conventional PET reconstruction were subsequently classified as consistent negative (3 negative ratings) using PET reconstruction with MR priors. Moreover, Fig. 6 showcases three subjects who transitioned from inconsistent negative to consistent positive (3 positive ratings) after applying the MR prior-based reconstruction method. In all these cases the agreement between trained readers was improved and for the 3 cases shown in Fig. 6, the diagnostic conclusion changed from amyloid negative to amyloid positive.

**Fig. 5.**
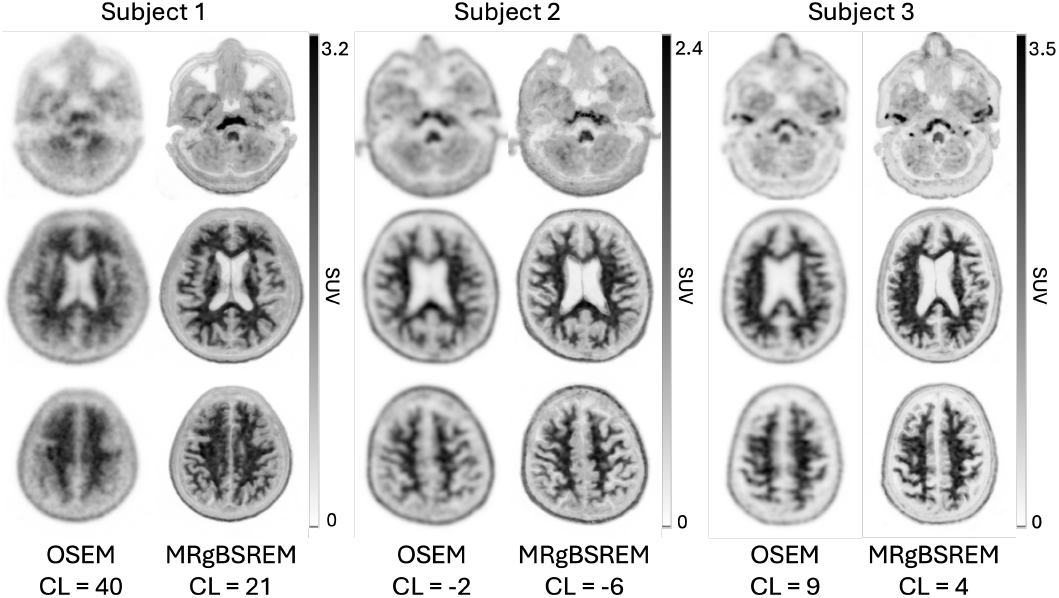
Shows three subjects initially classified as inconsistent negative (two negative, one positive rating) using OSEM PET reconstruction. Subsequent analysis using MRgBSREM PET reconstruction reclassified these subjects as consistent negative (three negative ratings).

**Fig. 6.**
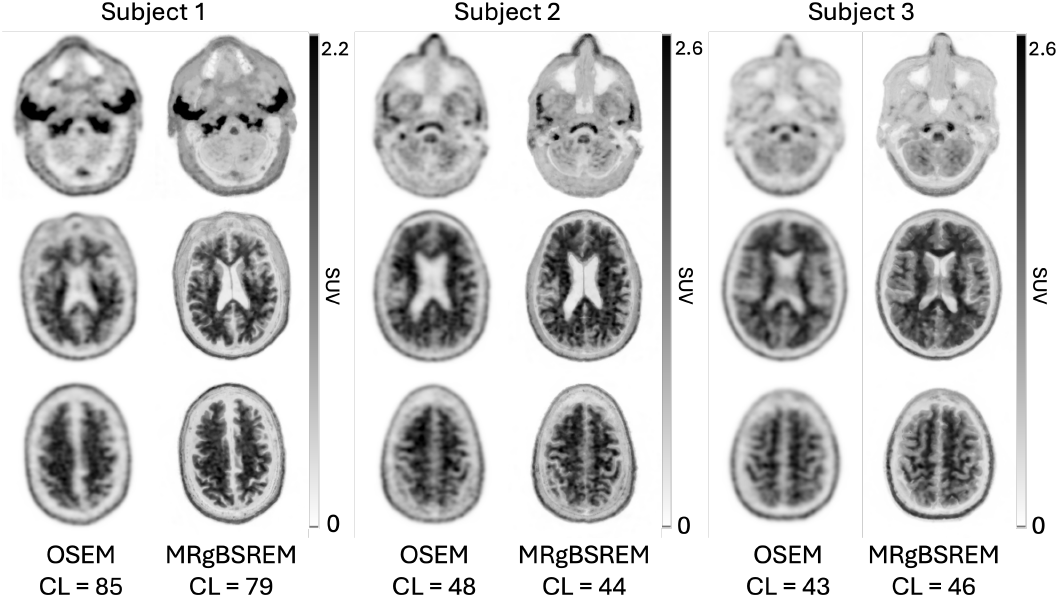
Shows three subjects initially classified as inconsistent negative (two negative, one positive rating) using OSEM PET reconstruction. Subsequent analysis using MRgBSREM PET reconstruction reclassified these subjects as consistent positive (three positive ratings).

Fig. 7 illustrates the distribution of CL derived from OSEM and MRgBSREM PET reconstruction methods for a subset of 106 subjects who were rated by our trained readers based on both OSEM and MRgBSREM PET images. Half of these subjects (53 subjects) were rated inconsistently using the conventional OSEM PET reconstruction method. The CL are divided into four groups based on their ratings: NNN (three negative readings), NNP (two negative and one positive reading), NPP (two positive and one negative reading), and PPP (three positive readings). Fig. 7(a) shows the distribution of CL measured by the OSEM PET reconstruction method, separated by their ratings based on the same method. Fig. 7(b) shows the same distribution of CL measured by the MRgBSREM PET reconstruction method, using the ratings based on PET images reconstructed by MRgBSREM. A comparison between Fig. 7(a) and Fig. 7(b) reveals that MRgBSREM has increased the agreement between readers and reduced the number of inconsistent cases (NNP or NPP groups). It also demonstrates that the inconsistent cases for the MRgBSREM method are clustered around the borderline between consistent negative and positive cases on the CL axis. In contrast, the inconsistent readings based on the OSEM method are more spread across the CL axis. Additionally, Fig. 7(b) shows that the number of consistently rated positive (PPP) cases has increased using the MRgBSREM PET reconstruction method, and these cases are more dispersed along the CL axis.

**Fig. 7.**
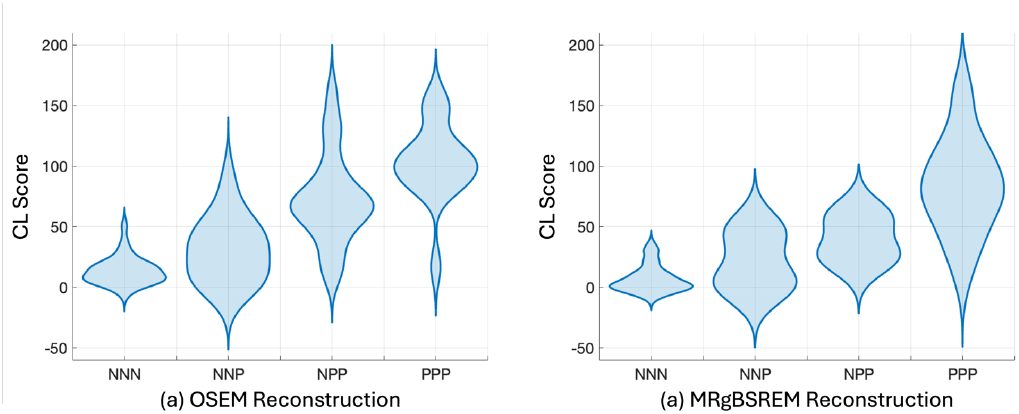
Shows the distribution of CL derived from OSEM and MRgBSREM methods for a subset of 106 subjects where half of them were rated inconsistently using OSEM method. The CL are divided into four groups based on their ratings: NNN (three negative readings), NNP (two negative and one positive reading), NPP (two positive and one negative reading), and PPP (three positive readings). A comparison between Figure 7(a) and Figure 7(b) reveals that the inconsistent cases (labeled by NNP and NPP) for the MRgBSREM method are clustered around the borderline between consistent negative (NNN) and positive (PPP) cases on the CL axis. In contrast, the inconsistent readings based on the OSEM method are more spread across the CL axis. Figure 7(b) shows that the number of consistently rated positive (PPP) cases has increased using the MRgBSREM PET reconstruction method, and these cases are more dispersed along the CL axis.

Fig. 8 illustrates the distribution of CL derived from OSEM and MRgBSREM PET reconstruction methods on the 106 rated subjects, with color-coding based on their rating category: NNN, NNP, NPP or PPP. Fig. 8(a) and 8(b) demonstrate a strong correlation (R2 = 0.99) between CL derived from the two methods. The color-coding highlights how some inconsistent ratings obtained using the OSEM method have changed into consistent ratings using the MRgBSREM method. The Bland-Altman plots shown in Fig. 8(c) and 8(d) reveal lower CL for negative cases (NNN or NNP groups) and higher CL for positive cases (NPP or PPP groups) when using the MRgBSREM method compared to the OSEM method. This difference is attributable to the spill-in and spill-over artifact reduction capabilities of the MRgBSREM method, which enables more accurate quantification of local FBB uptake.

**Fig. 8.**
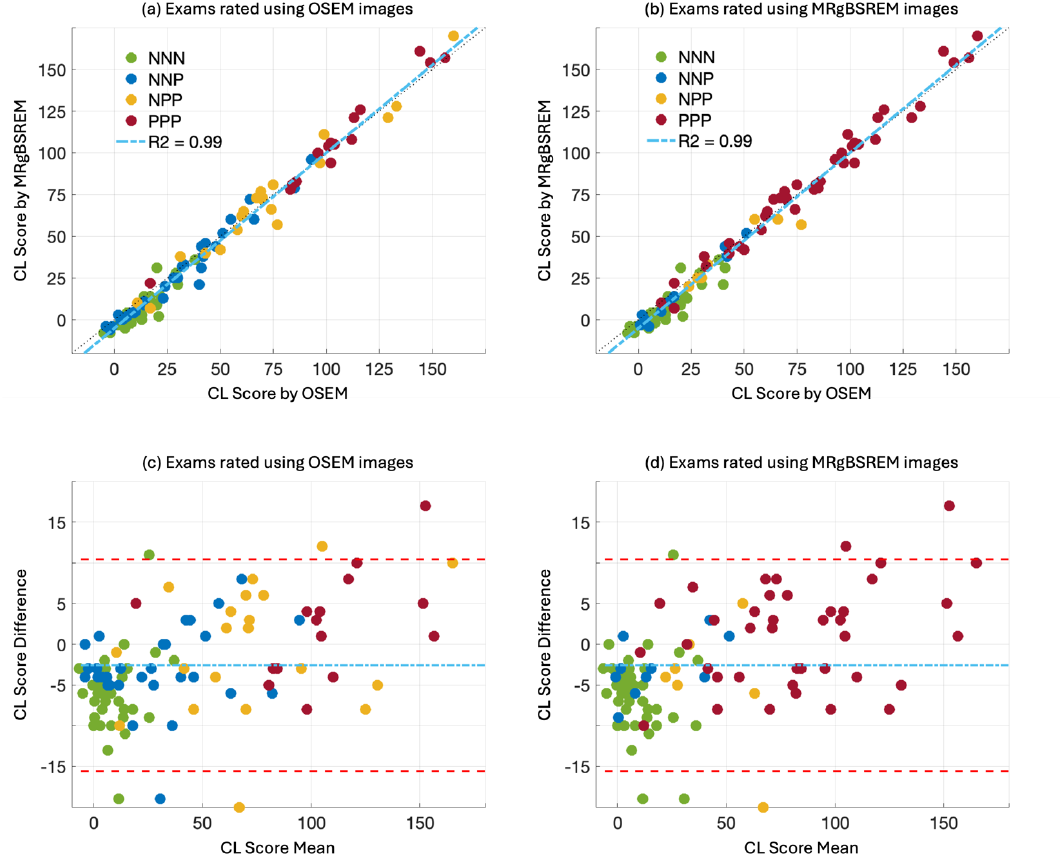
Illustrates the distribution of CL derived from OSEM and MRgBSREM methods, using a subset of 106 subjects. Each subject is color-coded based on their rating category: NNN (three negative readings), NNP (two negative and one positive reading), NPP (two positive and one negative reading), and PPP (three positive readings). The color-coding in Figures 8(a) and 8(b) highlights how some inconsistent ratings obtained using the OSEM method have changed into consistent ratings using the MRgBSREM method. The Bland-Altman plots shown in Figures 8(c) and 8(d) reveal lower CL for negative cases (NNN or NNP groups) and higher CL for positive cases (NPP or PPP groups) when using the MRgBSREM method compared to the OSEM method.

Table 1 presents a summary of the ratings for a subset of 106 subjects who were rated using PET images reconstructed with both OSEM and MRgBSREM methods. The total number of inconsistent cases (NNP and NPP groups) decreased significantly from 53 exams (50%) using the conventional OSEM PET reconstruction method to 20 exams (19%) using PET reconstruction with MR priors. By leveraging MR priors, 62% (33 exams) of the inconsistent cases were resolved, and 79% of these resolved cases (26 exams) were rated consistently positive, demonstrating the potential clinical impact of MR prior-based PET reconstruction.

**Table 1.**
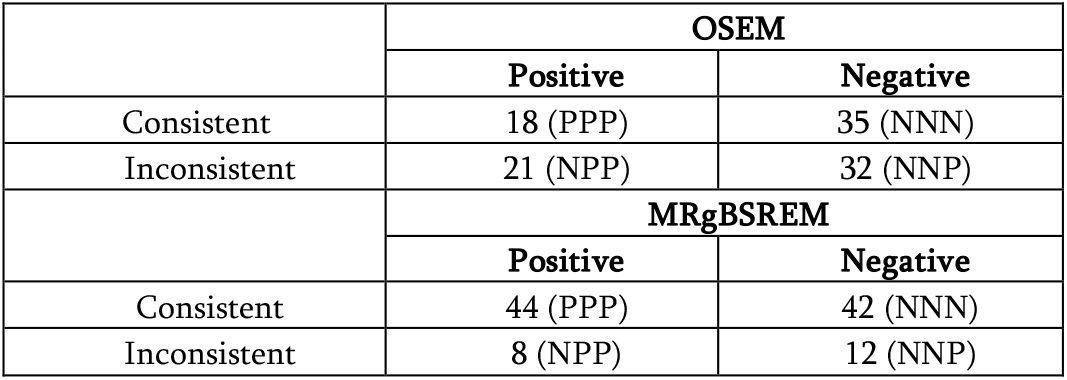
Number of ratings for consistent positive (three positive readings: PPP), inconsistent positive (two positive and one negative reading: NPP), inconsistent negative (two negative and one positive reading: NNP), and consistent negative (three negative readings: NNN) groups.

|  | OSEM |  |
| --- | --- | --- |
|  | Positive | Negative |
| Consistent | 18 (PPP) | 35 (NNN) |
| Inconsistent | 21 (NPP) | 32 (NNP) |
|  | MRgBSREM |  |
|  | Positive | Negative |
| Consistent | 44 (PPP) | 42 (NNN) |
| Inconsistent | 8 (NPP) | 12 (NNP) |

Fleiss’ kappa, a statistical measure to assess inter-rater agreement, was measured at 0.71 (95% CI: 0.70-0.73), for all 264 exams with ratings based on conventional OSEM PET reconstruction, indicating ‘Substantial’ agreement with a Z-score of 20.2. Measuring in the smaller cohort of 106 showed an expected lower Fleiss’ kappa of 0.31 (95% confidence interval: 0.28-0.34) for ratings based on the OSEM PET reconstruction method, indicating ‘Fair’ agreement with a Z-score of 5.5. By leveraging MR priors, PET reconstruction with MRgBSREM significantly improved inter-rater agreement, with Fleiss’ kappa increasing to 0.62 (95% confidence interval: 0.59-0.65), indicating ‘Substantial’ agreement. In the control group, the number of inconsistent ratings decreased marginally from 13 cases (50%) to 12 cases (46%) between the first and second readings of conventional PET images, and Fleiss’ kappa remained ‘Fair’ for both readings, with values of 0.3 and 0.37, respectively. The average intraclass correlation coefficient, a statistical measure to assess intra-rater agreement, was measured at 0.71 in the control group, which shows substantial agreement between the reader’s repeated ratings, suggesting a good level of consistency in their assessments.

## IV. Discussion

The MRgBSREM PET reconstruction method exploits the correlation between similar voxels found on MR images to penalize the cost function in an iterative PET reconstruction algorithm. By utilizing MR priors, MRgBSREM addresses the spill-in and spill-over artifacts on low resolution PET imagers. This method has been shown to improved reader agreement by providing higher image resolution and better image quality resulting in the enhanced gray-matter white-matter boundaries. This method also increased the differentiation in CLs between visually negative and visually positive scans, highlighting it’s utility for quantitative analyses.

As shown in [4], the scanner detector has an in-plane resolution of > 4 mm and a trans-axial resolution of > 6 mm. OSEM PET images were reconstructed using an in-plane Gaussian filter with a 3.5 mm FWHM cutoff spatial frequency and an axial FIR filter with coefficients [1, 4, 1]. These filters were chosen to minimize noise while preserving image resolution relative to the detector’s inherent resolution. Furthermore, the OSEM images were reconstructed from four 5-minute frames and subsequently co-registered to correct for subject large rigid motion.

Using MRgBSREM method for visual interpretation may require some adjustment in rating criteria as the gray matter is better defined, even in negative cases. To address this, a consensus was reached among the raters to adopt a revised criterion for MR-guided reconstructions. Instead of simply assessing the presence of cortical uptake as indicative of abnormality, the raters agreed to consider increased cortical uptake as the defining characteristic of abnormality, given the enhanced gray-white matter contrast afforded by MR-guided reconstructions, regardless of case positivity or negativity.

The agreement between reader for all 264 subjects was measured ‘Substantial’ with a Z-score of 20.2; however, it was measured ‘Fair’ with a Z-score of 5.5 on the subset of 106. This was expected as we intentionally selected half the cases with disagreement. However, MRgBSREM method was able to improve the readers’ agreement on this subset from “Fair” to ‘Substantial’ with a Z-score of 11.1.

One of the advantages of using PET images reconstructed with MRgBSREM method for visual rating of amyloid exams is that inconsistent ratings are no longer dispersed across cases with varying global SUVR values. The MRgBSREM method limited the inconsistent cases to those that are borderline and challenging to interpret. The enhanced resolution and image quality afforded by PET images reconstructed with MRgBSREM may pave the way for the development of new guidelines for visual interpretation of amyloid images, which could aid in clarifying these borderline cases and improving diagnostic accuracy.

As shown in Fig. 4, the SNR of PET images reconstructed with MRgBSREM using β=50 and β_m_=25 is comparable to OSEM. However, increasing β and β_m_ in MRgBSREM enhances SNR, though it also increases the influence of MR images on PET reconstruction [9].

Theoretically, the MRgBSREM method can be applied using various anatomical priors, including high-resolution MR or CT images. It can be also used for PET reconstruction on PET/CT scanners, where a co-registered non-simultaneous MR image can be utilized. Given that head motion is primarily rigid, accurate co-registration of non-simultaneous MR and CT images is feasible [14].

Several MR-guided PET reconstruction methods have been proposed [7, 15-17]. A key advantage of MRgBSREM, as described in [9], is its use of a PET seed image to identify areas where anatomical priors can be effectively applied. This decoupling approach helps mitigate artifacts caused by potential mismatches between anatomical images and the true activity distribution.

One of the limitations of the MRgBSREM method is its dependence on a high-resolution MR image to provide accurate priors for reconstructing a high-resolution PET image. While the MRgBSREM method incorporates a built-in PET motion correction algorithm [11] to address patient motion during the PET scan, it is important to note that the presence of motion artifacts on the MR images can limit the method’s ability to achieve the same level of high quality and high-resolution image reconstruction. However, it is crucial to emphasize that any motion artifacts present on the MR images will not directly introduce artifacts into the final reconstructed PET image as MR priors’ inclusion in MRgBSREM method is controlled by a PET seed image [9].

## Conclusion

PET reconstruction with MR priors can significantly improve the consistency of ratings among trained readers. Given the prevalence of inconsistent ratings in amyloid PET, methods that enhance the ability to distinguish intermediate amyloid levels could be valuable for the widespread adoption of this modality.

## Data Availability

All data produced in the present study are available upon reasonable request to the authors

## References

[1] Montine TJ, Phelps CH, Beach TG, Bigio EH, Cairns NJ, Dickson DW, et al. National Institute on Aging-Alzheimer’s Association guidelines for the neuropathologic assessment of Alzheimer’s disease: a practical approach. Acta Neuropathol. 2012;123:1–11.

[2] Fleisher AS, Pontecorvo MJ, Devous MD Sr, Lu M, Arora AK, Truocchio SP, et al. Positron emission tomography imaging with [18F]flortaucipir and postmortem assessment of Alzheimer disease neuropathologic changes. JAMA Neurol. 2020.

[3] Levin CS, Maramraju HS, Khalighi MM, Deller TW, Delso G, Jansen F. Design Features and Mutual Compatibility Studies of the Time-of-Flight PET Capable GE SIGNA PET/MR System. IEEE TMI. 2016; 35(8):1907–1914.

[4] Grant A, Deller TW, Khalighi MM, Maramraju SH, Delso G, Levin CS. NEMA NU 2-2012 performance studies for the SiPM-based ToF-PET component of the GE SIGNA PET/MR system, Med. Phys. 2016; 43:2334.

[5] Klunk WE, Koeppe RA, Price JC, et al. “The Centiloid Project: standardizing quantitative amyloid plaque estimation by PET,” Alzheimers Dement. 2015; 11:1–15. e154.

[6] Johns E, Vossler HA, Young CB, et al. “Florbetaben amyloid PET acquisition time: Influence on Centiloids and interpretation.” Alzheimer’s Dement. 2024; 20:5299–5310.

[7] Schramm G, Holler G, Rezaeiet A, et al. “Evaluation of Parallel Level Sets and Bowsher’s Method as Segmentation-Free Anatomical Priors for Time-of-Flight PET Reconstruction.” IEEE Trans Med Imaging 2018. 37(2), pp 590–603.

[8] Shepherd T., Schramm G., Vahle T., et al. “Impact of MR-guided PET Reconstruction on Seizure Foci Localization with FDG PET,” Journal of Nuclear Medicine May 2019, 60 (supplement 1) 392;

[9] Khalighi MM, Young CB, Spangler-Bickell MG, et al. “A Novel Method in PET Image Reconstruction Using MRI Anatomical Priors,” in IEEE Transaction on Radiation and Plasma Medical Sciences. 2025, doi: 10.1109/TRPMS.2025.3553409.

[10] Delso G, Wiesinger F, Sacolick LI, Kaushik SS, Shanbhag DD, Hüllner M., and Veit-Haibach P., “Clinical Evaluation of Zero-Echo-Time MR Imaging for the Segmentation of the Skull,” Journal of Nuclear Medicine March 2015, 56 (3) 417–422;

[11] Spangler-Bickell M., Hurley S., Pirasteh A., Perlman S., Deller T., McMillan A., “Evaluation of Data-Driven Rigid Motion Correction in Clinical Brain PET Imaging,” Journal of Nuclear Medicine, October 2022, 63 (10) 1604–1610.

[12] Khalighi MM, Deller T., Spangler-Bickell M., Wangerin, K. Holley D., Halbert K., Zeineh M., Zaharchuk G., Mormino E., Iagaru A., and Moseley M., “High Quality Isotropic Whole-body PET Imaging Using MR Priors,” J Nucl Med May 2020 vol. 61 no. supplement 1 1477

[13] Royse SK, Minhas DS, Lopresti BJ, et al. “Validation of amyloid PET positivity thresholds in centiloids: a multisite PET study approach,” Alzheimers Res Ther. 2021; 13:99. doi:10.1186/s13195-021-00836-1

[14] Tsai Y, Bousse A, Ahn A, et al. “Algorithms for Solving Misalignment Issues in Penalized PET/CT Reconstruction Using Anatomical Priors,” in Proc. IEEE Nuclear Science Symposium and Medical Imaging Conference Proceedings (NSS/MIC), 2018, pp 1–3.

[15] M. Ehrhardt, Markiewicz P, Liljeroth M, et al. “PET Reconstruction with an Anatomical MRI Prior using Parallel Level Sets.” IEEE Trans Med Imaging. 2016; 35(9), pp 2189–2199

[16] Mehranian A, Belzunce MA, Niccolini F, et al. “PET image reconstruction using multi-parametric anato-functional priors.” Phys. Med. Biol. 2017; 62, pp 5975–6007.

[17] Bland J, Mehranian A, Belzunceet MA, et al. “Intercomparison of MR-informed PET image reconstruction methods.” Med Phys. 2019; 46(11), pp 5055–5074.

